# Impact of a Mobile Application for Public-Private Tuberculosis Screening on Case Detection and Notification in Nigeria

**DOI:** 10.64898/2026.03.19.26348819

**Authors:** Bethrand Odume, Chidubem Ogbudebe, Micheal Sheshi, Iboro Gordon, Sabastine Wakdok, Elias Aniwada, Ademola Serrano, Njide Ndili, Tobias F. Rinke de Wit, Gloria P. Gómez-Pérez, Fati Murtala-Ibrahim, Obioma Chijioke-Akaniro, Emperor Ubochioma

## Abstract

**Background:** Tuberculosis remains a major public health concern in Nigeria, with significant diagnostic and treatment gaps, particularly within the public-private mix healthcare sector. To address these challenges, the Mobile Application for Tuberculosis Screening was introduced as a digital tool to strengthen TB screening, presumptive case identification, and notification across diverse healthcare settings.

**Methods:** A mixed-method study design was employed, integrating retrospective quantitative data analysis using a comparative analysis approach with qualitative thematic analysis. Quantitative data were drawn from the MATS database, routine facility records, and national TB surveillance systems, comparing pre- and post-MATS implementation periods. Key Informant Interviews and Focus Group Discussions were conducted with TB patients, health workers, and program managers across five geopolitical zones to explore user experiences, operational realities, and systemic barriers.

**Results:** Between 2021 and 2023, a total of 1,048,575 individuals were screened using MATS across 24 states, identifying 283,807 presumptive TB cases and a diagnostic yield of 16.9% (4,725). The most notable TB case gains were in the North-West and North-Central regions. Private-for-profit facilities and patent medicine vendors contributed over 60% of presumptive identifications, underscoring the importance of engaging non-state actors in TB control. Qualitative findings highlighted positive user experiences and improved data management but also revealed barriers to diagnostic follow-through, including logistical constraints, economic challenges, and health system inefficiencies.

**Conclusion:** MATS significantly improved the efficiency and quality of Nigeria’s private sector TB case detection and strengthened TB surveillance. While operational barriers remain, the tool demonstrates strong potential for further scale-up and integration into broader disease management frameworks. Future success will depend on sustained investment in digital infrastructure, strengthening referral and linkages, and patient support to optimise impact and advance national TB control goals.

## INTRODUCTION

Tuberculosis (TB) remains one of the leading causes of infectious disease-related morbidity and mortality globally, claiming over 1.3 million lives each year, despite being largely preventable and curable (1). The burden is disproportionately borne by low- and middle-income countries (LMICs), with sub-Saharan Africa accounting for a significant share of global TB incidence. Nigeria, Africa’s most populous country with over 220 million people, has persistently ranked among the top 10 countries globally with the highest TB burden (2). As of 2023, Nigeria ranked sixth in global TB incidence and first in Africa, with an estimated 452,000 new TB cases annually, nearly one-third of which remain undiagnosed or unreported (1,2).

Over the past few years, Nigeria has made considerable progress in scaling up TB case notifications, particularly through community-based interventions and public-private mix (PPM) strategies. Notably, TB case notifications increased from 138,591 in 2020 to 207,785 in 2021 and 285,561 in 2022, marking a 50% increase between 2020 and 2022 (1,3).

Despite these gains, a significant diagnostic and treatment gap persists in Nigeria’s TB response, with approximately 38% of TB cases still missing each year (3). Contributing factors include limited access to diagnostic services in rural and peri-urban areas, fragmentation between public and private providers, and challenges in real-time surveillance and data capture (2,4,5). Nigeria’s TB surveillance system continues to struggle with incomplete reporting and weak case-finding mechanisms (6,7), underscoring the urgent need to strengthen access to high-quality TB screening, diagnosis, and treatment services.

One promising avenue for addressing these gaps is the application of digital health technologies. Digital tools such as mobile phone applications, Short Message Service (SMS) platforms, and wearable devices are increasingly recognised for their role in improving health outcomes, particularly in LMICs (8–17). These have also proven valuable during the COVID-19 pandemic, when remote patient support and digital surveillance became indispensable (18). In the context of TB, digital technologies have been used to support disease screening, case notification, treatment adherence, patient follow-up, and real-time communication between patients and healthcare providers (19,20).

In Nigeria, private healthcare providers are critical players in the healthcare delivery ecosystem, contributing up to 70% of health services (21,22). Strengthening their role in TB control through mandatory notification and improved digital surveillance could significantly bolster national TB response efforts (23,24). However, weak notification systems in the private sector have led to underestimation of TB burden, misallocation of resources, and suboptimal implementation of control strategies (7,22). Evidence from Nigeria suggests that effective engagement of private providers in TB control, facilitated by digital technologies, can increase case detection and benefit both patients and providers (24,26).

Given the widespread use of mobile phones in Africa, with mobile penetration projected to reach 84% by the end of 2025, mobile-based health interventions present a scalable and cost-effective opportunity to enhance TB control in Nigeria (27,28). These tools have already shown promise in improving medication adherence, disease tracking, and healthcare access in low-resource settings burdened by high infectious disease prevalence (27,28). Additionally, mobile health technologies have been leveraged to strengthen private-public mix (PPM) models for TB care, although evidence on their effectiveness and feasibility remains limited (29,30). Due to this lack of robust data, the World Health Organisation (WHO) currently offers only a conditional recommendation for the use of digital tools in TB treatment (19).

The current study assesses the utilisation, effectiveness, and feasibility of digital technologies for TB screening and case notification within the PPM context in Nigeria. By exploring how these interventions can bridge the TB detection and treatment gap, this research aims to contribute to the growing body of knowledge that supports the integration of digital innovations in national TB control strategies. The aim of this study was to assess the impact of MATS on TB case notification rates in selected states in Nigeria. The study also seeks to understand the usability, accessibility, and effectiveness of MATS in PPM healthcare settings in Nigeria. Specific objectives were: 1. To determine how effectively the MATS application has improved TB case detection, presumptive case identification, and notification in PPM healthcare facilities; 2. To determine the contribution of MATS to national TB control goals, such as increasing TB case detection and reducing the burden of TB in Nigeria, 3. To compare TB case notification rates before and after the implementation of MATS to evaluate its direct impact, 4. To assess MATS integration with and contribution to the national TB surveillance system, and how this integration enhances TB control efforts in Nigeria. 5. To provide recommendations for scaling up the application, integrating it with other health interventions, or modifying it for better effectiveness and 6. To evaluate the experience of individuals identified as presumptive TB cases through MATS, including the challenges and barriers faced in accessing testing and treatment.

## METHODS

### Study Setting

The study was conducted across selected private-sector health facilities in Nigeria, spanning five of the six geopolitical zones: North-West, North-Central, South-West, South-East, and South-South; none of the North-East states implemented MATS due to persistent insecurity and insurgency. Nigeria comprises 36 states and the Federal Capital Territory (FCT), with significant diversity in geography, culture, religion, and health service delivery structures. This broad geographical distribution was purposively chosen to reflect the heterogeneous nature of Nigeria’s health system and to enhance the generalizability of findings.

Health service delivery in Nigeria is stratified across three tiers of government: the federal government manages tertiary health institutions; state governments oversee secondary health facilities; and local governments operate primary health centres. Tuberculosis services are delivered at all levels of care, with coordinated oversight from the National Tuberculosis and Leprosy Control Programme (NTBLCP).

### Study Design

A mixed-method design was employed. For the quantitative component, we used a comparative analysis approach to evaluate the impact of MATS on TB case notifications. The qualitative component involved thematic analysis of key in-depth interviews (KIIs) and focus group discussions (FGDs) to explore stakeholder experiences and the implementation dynamics of MATS.

The study employed a retrospective data collection of TB cascade data from before (2017 – 2019) and after (2021 – 2023) the implementation of MATS within intervention facilities.

### Study Population and Sample

The quantitative population comprised all presumptive TB cases recorded in the MATS app across 24 states during the study period. A total population study design was adopted, involving all eligible presumptive TB cases with complete TB cascade data during the pre- and post-MATS phases.

For the qualitative component, we purposively selected key stakeholders engaged with MATS, including government officials, health workers, implementing partners, and patients. A total of 60 Key Informant Interviews (KIIs) were conducted, with 12 participants per state, covering: 2 State TB and Leprosy Supervisors (TBLS), 2 Directly Observed Therapy (DOT) Officers, 1 State TB Programme Manager, 1 State TB Program Monitoring & Evaluation focal person, 2 Implementing Partner staff, 2 MATS clients, and 2 Linkage Coordinators. Additionally, six FGDs covering the geopolitical zones were held, with a minimum of eight participants in each group.

### Data Sources and Collection

i. (i) MATS tool The Mobile Application for Tuberculosis Screening (MATS) is a digital screening and referral application used by trained providers in participating sector facilities to identify individuals with presumptive TB and support linkage to diagnostic testing. During routine consultations, providers administer a structured set of screening questions within the application, consistent with the national TB symptom screening approach and an embedded clinical decision algorithm. Client responses are processed by the in-app algorithm, which classifies individuals as “presumptive TB” when predefined criteria are met. Once classified, the application prompts referral/linkage for TB diagnostic evaluation and enables recording of subsequent cascade events (e.g., diagnostic testing performed, test results, and confirmed TB notification where available). For this study, a presumptive TB case was defined as any individual recorded in the MATS database as presumptive TB based on the application algorithm during the study period.
ii. Quantitative data sources The quantitative data for this study were obtained from three primary sources: the MATS application database, hosted by the NTBLCP in collaboration with the Institute of Human Virology Nigeria (IHVN); routine health facility records; and national TB surveillance data maintained by the NTBLCP. These data sources provided comprehensive and complementary insights into the TB screening and notification processes across both public and private healthcare settings. Data were extracted retrospectively to enable a comparative analysis of trends and outcomes across two distinct periods: the pre-implementation (baseline) period, which served as the control phase, and the post-implementation (intervention) period, during which the MATS application was actively deployed. This approach allowed for a robust assessment of the impact of MATS on key TB indicators, including presumptive case identification, diagnostic evaluation, and confirmed case notifications. The use of routinely collected programme data also ensured the analysis was grounded in real-world service delivery conditions.
iii. Qualitative data collection To complement the quantitative findings and gain deeper insight into the implementation and operational dynamics of MATS, qualitative data were collected through in-depth interviews (IDIs) and focus group discussions (FGDs). Trained qualitative researchers led this component of the study, employing semi-structured interview guides designed to elicit detailed, open-ended responses from participants. These guides were tailored to explore perceptions, experiences, challenges, and enabling factors related to TB screening and notification, particularly related to the use of the MATS application.

Interviews were conducted with a diverse range of stakeholders, including frontline healthcare workers, linkage coordinators, facility managers, TB program officers, and community health influencers involved in MATS implementation. FGDs were also held with groups of health workers and patients to capture shared experiences and perspectives.

All interviews and FGDs were audio-recorded with prior informed consent from participants to ensure accuracy and transparency. The recordings were subsequently transcribed verbatim, and where necessary, translated into English to maintain fidelity to participants’ original expressions. In addition to recorded transcripts, researchers also documented detailed field notes and non-verbal observations during interviews and site visits. These supplementary materials were used to enrich the contextual understanding of the data, allowing for a more nuanced interpretation of the themes that emerged during analysis.

### Ethical Considerations

The study protocol received ethical approval from the National Health Research Ethics Committee (NHREC) of Nigeria (NHREC/01/01/2007-15/09/2021). For the quantitative component, all data were de-identified prior to analysis. For the qualitative component, informed consent was obtained from all participants. Participation was voluntary, and anonymity and confidentiality were strictly maintained with only initials used for the purposes of analysis.

### Data Analysis

1. Quantitative Analysis To estimate the causal impact of MATS on TB case notifications, we used the comparative analysis approach. This approach compared changes in TB indicators over time between health facilities that implemented MATS (intervention group) and comparable facilities that did not (control group). Primary outcomes analysed included (i) number of presumptive TB cases identified, (ii) proportion tested for TB, and (iii) number of confirmed TB diagnoses (all forms). Subgroup analyses were conducted by region (geopolitical zone), urban-rural location, and type of facility. Statistical analysis was conducted using STATA 17, with significance set at *p*⍰< ⍰0.05. Descriptive statistics were used to explore provider engagement with MATS (e.g., screening frequency, usage rates).
2. Qualitative Analysis The qualitative data were analysed using NVivo 14, a software platform for managing and coding qualitative data. All interview and focus group transcripts were subjected to a rigorous double-coding process by two independent analysts to enhance the reliability and consistency of the findings. This approach ensured intercoder reliability and minimised bias in data interpretation. The analysis followed Braun and Clarke’s six-step thematic analysis framework: familiarisation with the data; generating initial codes; searching for themes; reviewing themes; defining and naming themes; and producing the report. (31)

The themes that emerged from the analysis were developed inductively, allowing the data to speak for itself and reflect the lived realities of stakeholders involved in MATS implementation. This inductive process uncovered rich contextual insights into several key dimensions of the project. These included user experiences with the MATS application— particularly in terms of ease of use and perceived benefits, as well as barriers to implementation such as poor connectivity, increased workload, and resistance to change from some users. The analysis also highlighted how MATS was being integrated into routine TB surveillance and reporting systems, providing a clearer picture of its operational fit within existing workflows.

Furthermore, participants offered concrete recommendations for the scale-up and sustainability of MATS, drawing on their practical experience to propose solutions to enhance its future effectiveness. The themes were ultimately organised around the study’s specific objectives, with particular emphasis on objectives four through six, which focused on integration, scale-up, and user experience. This alignment ensured that the qualitative findings directly informed the evaluation of MATS within the broader goals of the study.

## RESULTS

### General characteristics of the MATS participants

A total of 283,807 presumptive TB cases were recorded across five geopolitical zones (no state in the North-East implemented MATS during the study period due to insecurity and insurgency-related factors) in Nigeria. Overall, females accounted for a slightly higher proportion (51.2%) than males (48.8%). Regional differences were notable: the South-East had the highest proportion of female cases (54.3%), while the North-West had the highest proportion of male cases (58.8%). See Table 1.

**Table 1:**
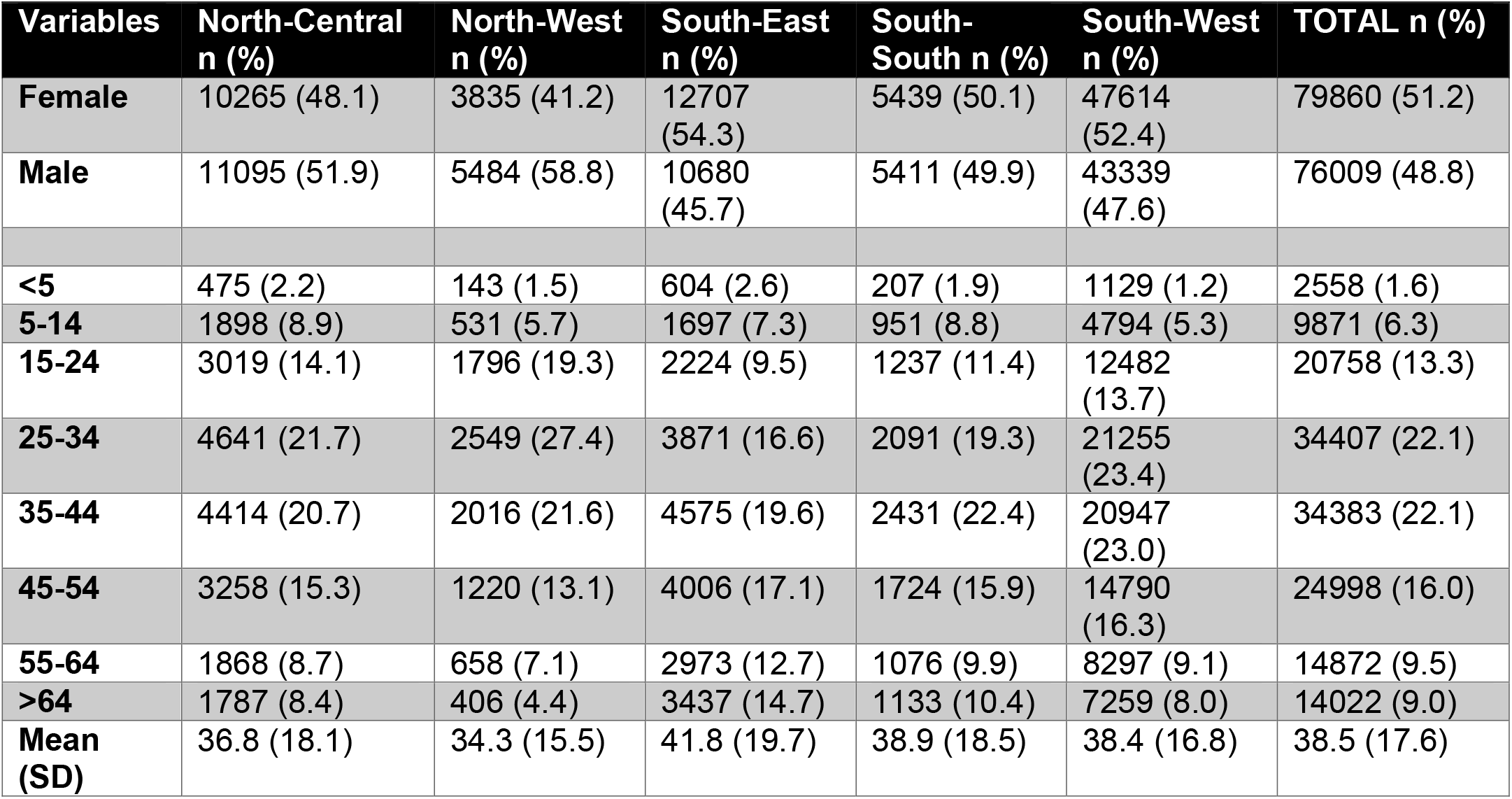
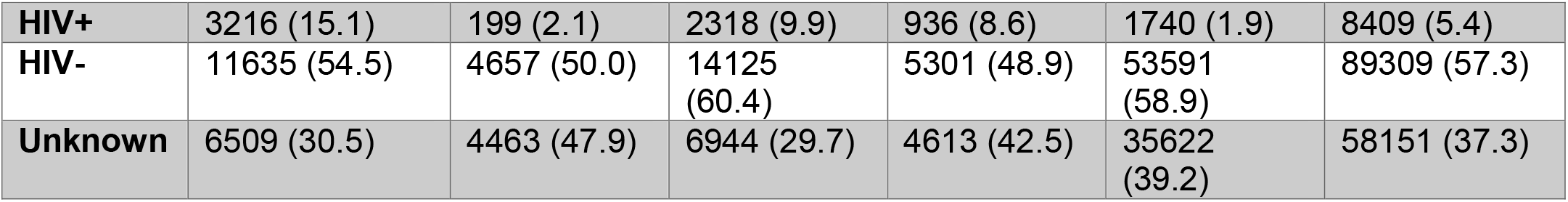
Regional characteristics of 155,869 MATS-identified presumptive TB cases referred for further evaluation.

Most presumptive TB cases were adults aged 25–44 years, making up over 44% of the total, with a national mean age of 38.5 years. The South-East had the highest average age (41.8 years), and the North-West the lowest (34.3 years). Children under five represent a small fraction of cases across all regions.

HIV co-infection varied significantly by region. Overall, 5.4% of presumptive TB cases were HIV-positive, 57.3% were HIV-negative, and 37.3% had unknown status. The North-Central zone had the highest HIV positivity rate (15.1%), while the North-West had the lowest (2.1%), although nearly half of the North-West cases had unknown HIV status.

#### Objective 1: Determine how effectively the MATS application has improved TB case detection, presumptive case identification, and notification in public-private mix healthcare facilities

Figure 1 presents the MATS screening cascade, illustrating the progressive drop-off from initial TB screening to confirmed diagnosis. A total of 1,048,575 individuals were screened using the MATS application across the selected facilities during the intervention period. Out of these, 283,807 individuals were identified as presumptive TB cases, reflecting the application’s capacity to rapidly triage high-risk individuals from the general screened population. This is a substantial yield that underscores the effectiveness of the digital tool in enhancing presumptive case identification, a critical first step in the TB diagnostic pathway.

**Figure 1.**
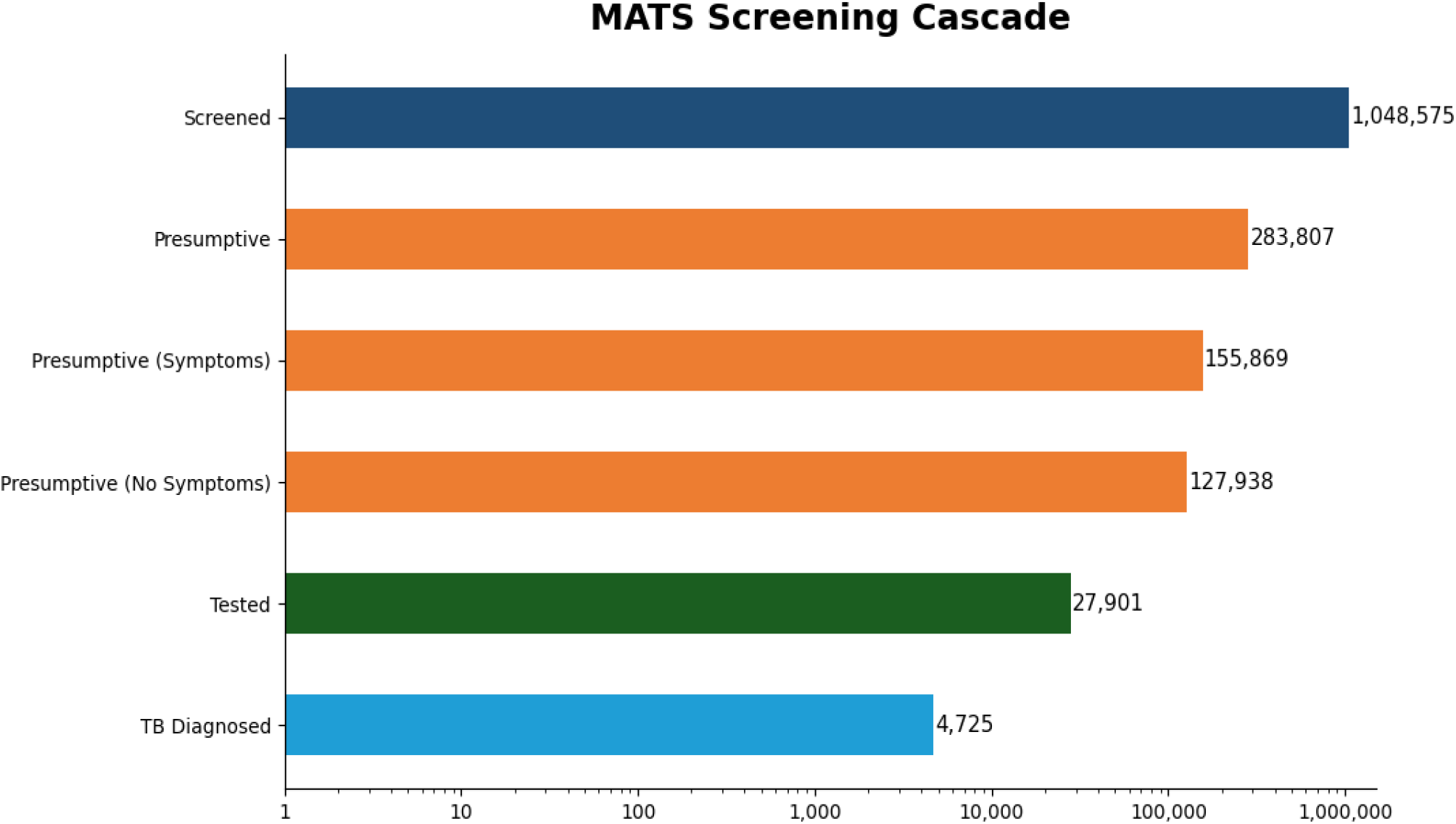
MATS Screening Cascade.

Following symptom-based screening within MATS, 127,938 records initially flagged as presumptive were excluded because they had zero TB-compatible symptoms, leaving 155,869 symptom-positive presumptive cases for cascade assessment. Of these, 27,901 (17.9%) proceeded to diagnostic evaluation. This drop-off is markedly higher than in the public sector (see also Table 2) pointing towards logistical, operational, and economic barriers, such as costs borne by patients (e.g., transport and time) that limited their ability to return for testing.

**Table 2.** Distribution of Presumptive and Actual TB Diagnosis Before and During MATS by Region and Total.

| Region | Period | Presumptive TB cases | Tested for TB (n) | Tested for TB (%) | Actual TB cases | % Diagnosed |
| --- | --- | --- | --- | --- | --- | --- |
| North-Central | Before | 76,235 | 74,826 | 98.2 | 4,439 | 5.9 |
|  | During | 42,383 | 307 | 0.7 | 98 | 31.9 |
| North-West | Before | 18,761 | 18,730 | 99.8 | 1,302 | 7.0 |
|  | During | 17,887 | 751 | 4.2 | 334 | 44.5 |
| South-East | Before | 51,786 | 51,133 | 98.7 | 2,696 | 5.3 |
|  | During | 39,262 | 7,512 | 19.1 | 1,826 | 24.3 |
| South-South | Before | 37,672 | 36,599 | 97.2 | 2,191 | 6.0 |
|  | During | 19,864 | 1,836 | 9.2 | 322 | 17.5 |
| South-West | Before | 54,536 | 52,358 | 96.0 | 4,621 | 8.8 |
|  | During | 164,411 | 17,495 | 10.6 | 2,145 | 12.3 |
| TOTAL | Before | 238,990 | 233,646 | 97.8 | 15,249 | 6.5 |
|  | During | 283,807 | 27,901 | 9.8 | 4,725 | 16.9 |

**Note:** *Before=mainly public facilities, after=public and private facilities. Public data collected from routine facility TB registers and NTBLCP surveillance/reporting databases*

**Table 3.**
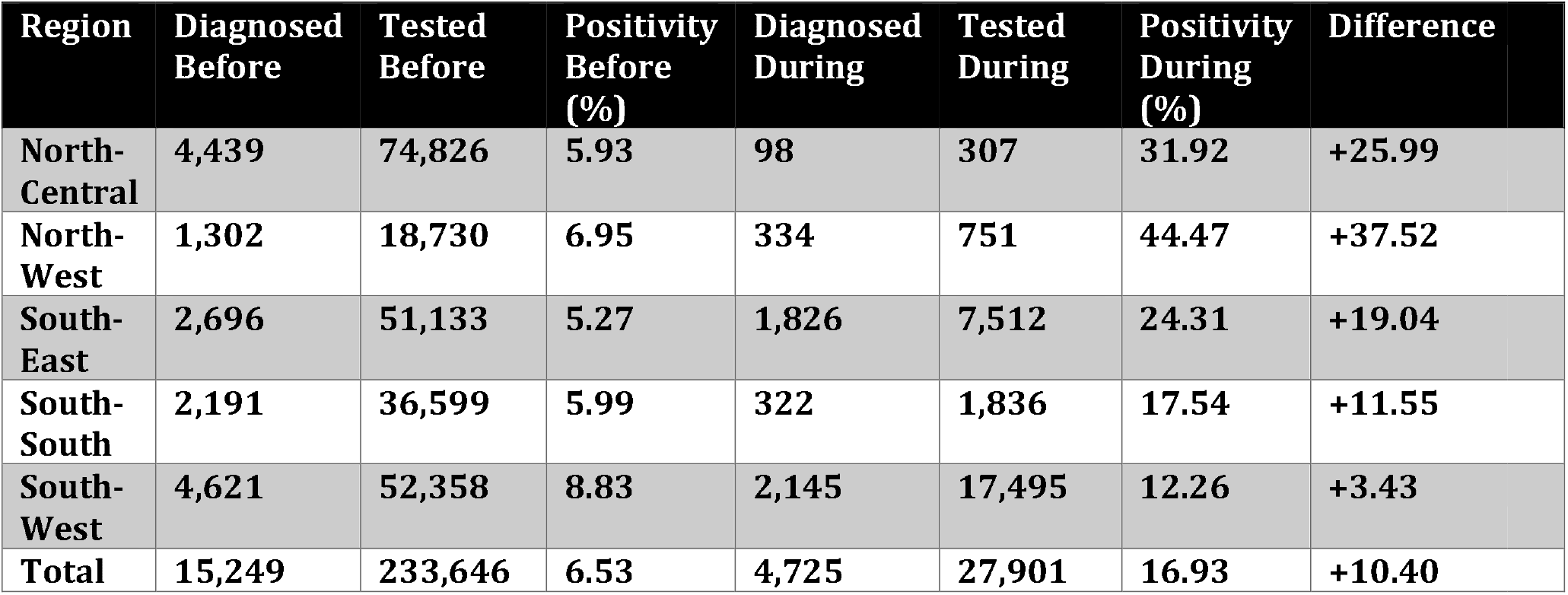
Regional positivity rate before and during MATS by region.

Among those who were tested for TB, 4,725 individuals were confirmed to have TB, representing a diagnostic yield of 16.9% (4,275/27,901). This is a strong diagnostic performance, particularly given that the pre-MATS baseline yield was markedly lower. The high positivity rate suggests that MATS effectively improved the sensitivity of case detection by enhancing the ability to flag high-risk individuals for diagnostic follow-up, thereby contributing to increased confirmed case notifications.

*Figure 2* provides further insight into where presumptive TB cases were being identified across different facility types. Notably, private-for-profit facilities accounted for 34.1% of all presumptive cases, followed closely by proprietary patent medicine vendors (PPMVs) at 28.4%. Together, these two groups contributed over 62% of all presumptive identifications, confirming the crucial role of the private sector in TB case finding within Nigeria’s pluralistic health system. Public health centres contributed 22.6%, while faith-based organizations and other facility types made smaller contributions (10.6% and 4.3%, respectively). This distribution reinforces the importance of integrating the private sector, often the first point of contact for many Nigerians, into the national TB response.

**Figure 2.**
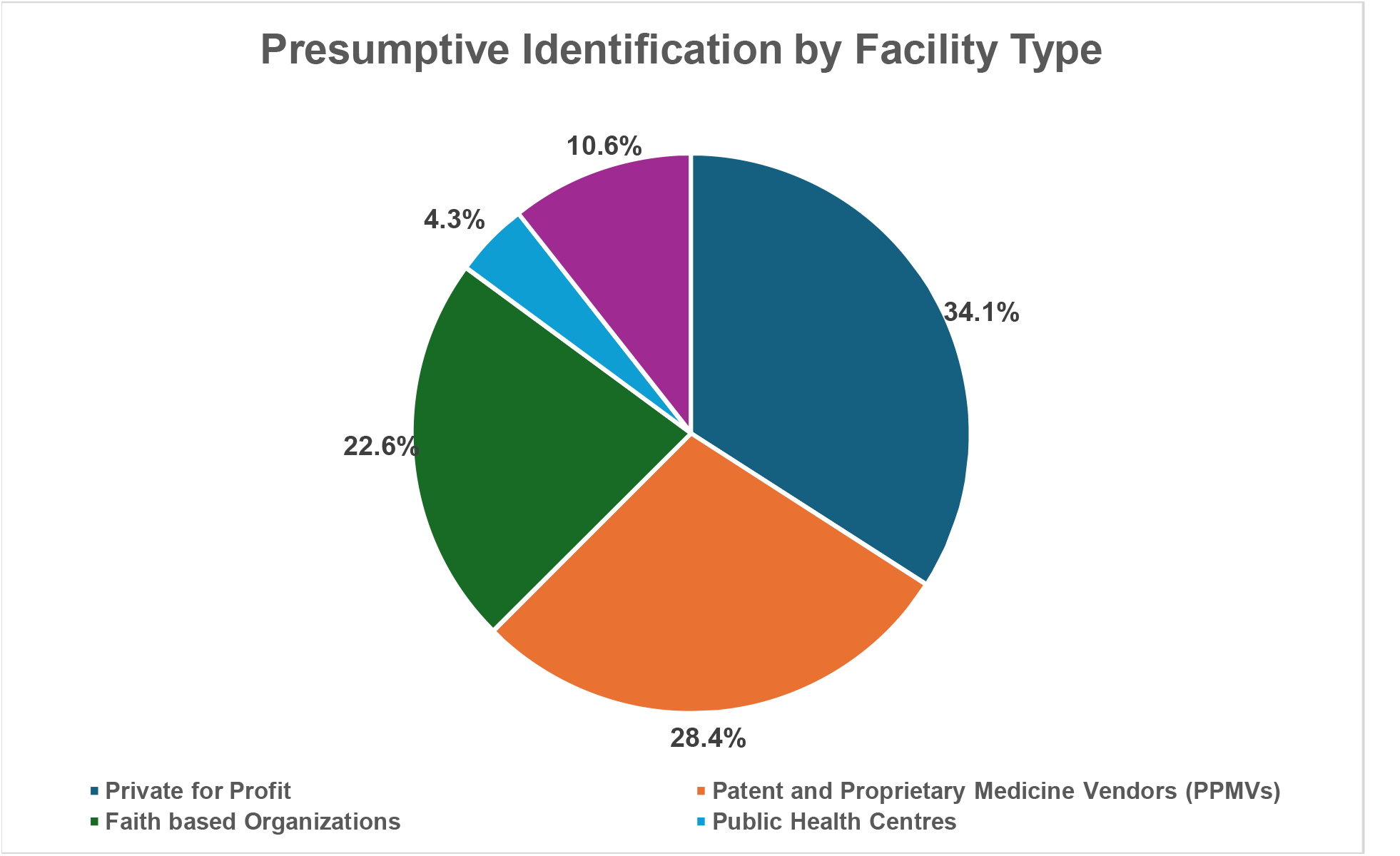
Presumptive Identification by Facility Type.

#### Objective 2: Evaluate the contribution of MATS to national TB control goals, particularly in increasing TB case detection and reducing the overall TB burden in Nigeria

In the pre-MATS period, 238,990 presumptive TB cases were identified, of which 233,646 (97.8%) underwent diagnostic evaluation (Table 2). This is typical for public sector TB care, in which according to guidelines all presumptive TB cases should be tested. From these, 15,249 individuals (6.5%) were diagnosed with TB.

During the MATS implementation period, the number of presumptive TB cases identified increased to 283,807—an additional contribution of 44,817 from the private sector (Table 2). Of the total presumptive cases 127,938 had no TB-compatible symptoms and were therefore deliberately excluded from diagnostic evaluation, leaving 155,869 symptomatic presumptive cases eligible for testing of which actually 17.9% did (27,901/155,869). This is a markedly lower testing rate than in the public sector (Table 2). However, the TB positivity yield among those who did get tested nearly tripled to 16.9% (4,725 cases), suggesting improved targeting of high-risk individuals and increased efficiency in case identification by MATS.

Regionally disaggregated analysis reveals further insights. The most change occurred in the South-West, where the number of presumptive TB cases surged from 54,536 before MATS to 164,411 during implementation. This significant increase highlights MATS’s ability to scale reach and coverage, especially in densely populated or underserved areas. Moreover, TB diagnostic yields improved markedly across all five regions, as seen in *Figure 3*. In the North-West, diagnostic yield rose from 7.0% to 44.5%; in the North-Central region, from 5.9% to 31.9%; and in the South-East, from 5.3% to 24.3%. Even in regions where absolute numbers of Presumptive declined (e.g., North-Central and South-South), the positivity rate rose sharply, reflecting enhanced precision in presumptive identification.

**Figure 3.**
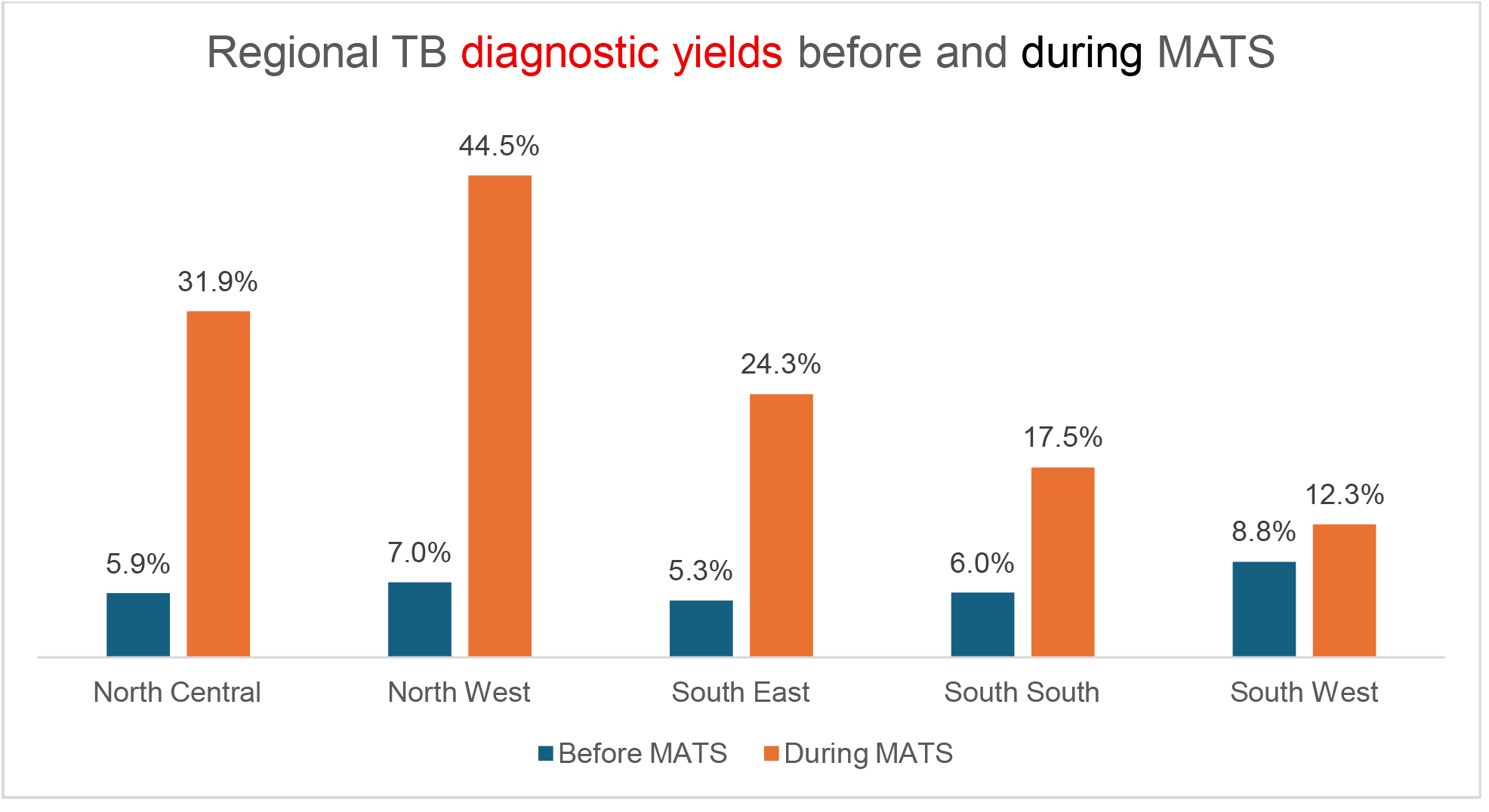
Regional TB Positivity Rates Before and During MATS.

**Figure 4.**
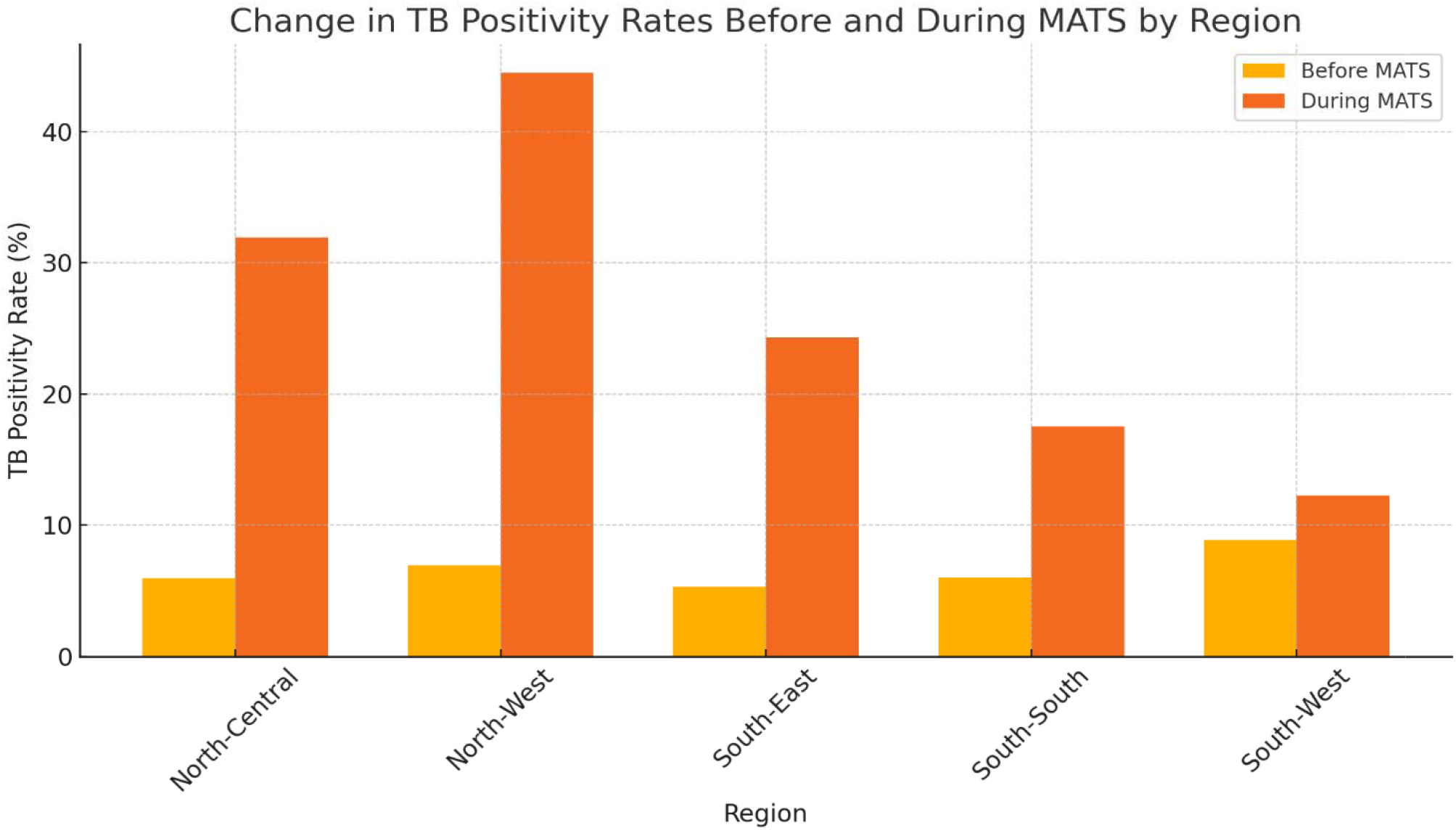
Change in TB positivity rates before and during MATS by region.

#### Objective 3: Compare TB case notification rates before and after the implementation of MATS, using appropriate statistical methods to estimate its direct impact

To assess the direct impact of the MATS application on TB case notification rates, a comparative analysis approach was applied, comparing TB diagnosis trends before and during the intervention. The analysis relied on aggregate TB program data from PPM facilities between 2017–2019 (pre-MATS) and MATS-generated data from 2021–2023 (during implementation). The pre-MATS cascade estimates were derived from routinely collected programme data (routine facility TB registers and NTBLCP surveillance/reporting databases) for the same facilities/period.

In the pre-MATS period, a total of 15,249 TB cases were diagnosed from 233,646 presumptive cases tested, representing a diagnostic yield of 6.5%. In contrast, during the MATS implementation period, 4,725 TB cases were diagnosed from 27,901 individuals tested, corresponding to a much higher diagnostic yield of 16.9%.

Despite the reduction in the absolute number of presumptive tested during the MATS period, the increase in diagnostic yield reflects a more targeted and efficient screening process, suggesting that the MATS application improved the identification of individuals with true TB.

To further quantify impact, we compared each region’s notification rates before and after MATS using the formula: *Positivity Rate (%) =Tested/TB Diagnosed × 100*

These results show an increase in detected TB positivity rates across all five geopolitical zones, with the most pronounced improvements observed in the North-West (+37.52 percentage points) and North-Central (+25.99 percentage points). These gains demonstrate the MATS platform’s effectiveness in improving the precision of TB case detection, especially in areas with historically lower diagnostic coverage.

#### Objective 4: Integration with TB Surveillance and Program Delivery

The qualitative findings from the KIIs conducted with clients, TBLS, DOT Screening Officers, and Healthcare Workers (HCWs) identified several salient themes regarding the effectiveness and implementation experience of MATS. The main themes identified included awareness and perception of MATS, benefits and positive experiences, challenges encountered, integration with health interventions, barriers influencing patient linkage, and recommendations for improvement.

Thematic analysis revealed that healthcare workers, particularly TBLS, DOT screening officers, and facility-based health staff, perceived MATS as a valuable digital innovation that enhances TB surveillance and control. Most respondents recognised its utility in improving data documentation, case tracking, and reporting accuracy, thereby supporting the broader TB program.

A DOT screening officer shared, *“The MATS app makes our work easier when it comes to documenting patient details”* (OY SO 1), emphasising the application’s role in simplifying surveillance tasks. Similarly, a healthcare worker stated, *“The MATS App is all about information… It’s a good thing to use”* (AK SO 1), pointing to the digital platform’s contribution to efficient information flow and real-time data access.

However, challenges such as poor internet connectivity, device limitations, and inconsistent uploads affected full integration. These operational issues at the frontlines often hindered seamless data flow into national systems. One officer expressed frustration with these barriers, noting, *“My experience with MATS implementation is all about the network issue… it is too dull”* (AK SO 2).

While the integration with the national TB surveillance system is promising, its full potential is currently limited by systemic infrastructural and connectivity issues, especially in rural or underserved areas. Despite these constraints, health workers’ overall positive perception of MATS and their active use of the application reflect its growing role as a surveillance-enabling tool in Nigeria’s TB control ecosystem.

#### Objective 5: Recommendations for Scale-Up and Integration

Respondents offered nuanced insights into how MATS could be scaled or adapted for broader health applications. While awareness of current integration with other health interventions was limited among most participants, a few TBLS and HCWs noted its potential for integration, particularly with HIV/AIDS programs. As one TBLS noted, *“I think there is integration because every TB case must be tested for HIV, and some cases come up with co-infection”* (PL TBLS 1). This reflects an opportunity to build a more holistic digital health ecosystem by leveraging MATS not only for TB but also for co-morbidities commonly seen in the same patient population.

Participants across all respondent groups also provided actionable recommendations for scaling and improving the application. These included the need for improved infrastructure, such as ensuring stable internet connectivity and providing dedicated smartphones for health workers using MATS. Additionally, capacity building was emphasised, specifically regular training and retraining of frontline staff to promote proficient and sustained use of the platform.

Financial and incentive support emerged as another critical enabler, with calls for monthly data subscriptions for health workers and transportation incentives for patients to enhance service uptake and continuity of care. Participants also recommended enhancing application functionality, such as increasing offline storage capacity, simplifying data entry processes, reducing the length of questionnaires, and enabling offline uploads to accommodate low-connectivity environments.

Community awareness creation was also identified as essential for scaling up MATS and increasing engagement. Respondents called for continuous media campaigns and public sensitisation, emphasising that TB testing and treatment are free to improve demand and reduce stigma-related delays in care-seeking.

Together, these recommendations speak to the technical, operational, and social elements required for the successful expansion and integration of MATS. They highlight the importance of addressing frontline realities while also envisioning the platform’s role in broader, more interconnected public health strategies

#### Objective 6: Client Experiences and Access Barriers

Clients identified through MATS shared generally positive experiences during the initial screening phase, describing the app-based interactions as efficient and user-friendly. One client remarked, *“I think it is easy and seamless and also less time-consuming…”* (KD CL 1), while another reflected on the structured nature of the questions and interactions with health workers as helpful for timely care. However, patient narratives also revealed critical barriers that hindered access to diagnostic and treatment services after initial identification. Several patients described economic and logistical burdens, such as transport costs and long travel distances to diagnostic centres. Stigma and fear of discrimination emerged as powerful deterrents. A client explained, *“I didn’t come for the treatment immediately because I was scared of discrimination…”* (PLP002), underlining the persistent social barriers affecting TB service uptake.

Others faced structural health system constraints. Long waiting times at facilities discouraged follow-through, with one participant recounting, *“I waited for more than four hours so I had to leave”* (AkP M004). These delays, coupled with staff attitudes and cultural beliefs, further disrupted the continuum of care. Awareness of MATS among clients was also limited. Most were introduced to the application only at the point of care, during facility visits or community outreach. A client explained, *“I got to know about it when I went to the hospital… the lady said it is the MATS app…that was how I got to know about the app”* (AN CL 1), suggesting that greater public sensitisation may be needed for broader engagement.

Collectively, these findings underscore that while MATS improved early engagement and presumptive identification, several downstream barriers, mostly systemic and socio-economic, still inhibit complete linkage to care. Addressing these challenges is essential to ensuring that the benefits of digital innovation translate into real-world health outcomes.

## DISCUSSION

This study set out to evaluate the impact of MATS on TB case detection, surveillance integration, and user experience within PPM healthcare settings in Nigeria. By employing comparative quantitative analysis with thematic synthesis of qualitative insights, the study provides compelling evidence that MATS contributed meaningfully to national TB control goals through improved targeting of private healthcare sector high-risk individuals, expanded case-finding reach, and strengthened surveillance responsiveness (1,2,3,4,9).

A notable finding was the substantial improvement in diagnostic yield following MATS implementation, which rose from 6.53% pre-MATS to 16.93%. This occurred despite a significant decline in the proportion of presumptive TB cases tested. Such a shift signals a critical reorientation from volume-based to quality-focused screening, ensuring that diagnostic efforts are better targeted toward individuals most likely to have TB (1,3,5). MATS’ higher diagnostic yield could be explained by its TB-tailored clinical algorithm comprising a set of questions designed to increase the likelihood of detecting presumptive TB cases. For example, it assesses not only cough and its duration, but also other symptoms such as night sweats, fever, and weight loss. In addition, early in the screening sequence, individuals are asked about HIV status, a known risk factor for TB, among others. This transition towards targeted diagnostic efforts is in line with WHO’s strategy for maximising efficiency in high-burden, resource-limited settings (1,5). The MATS screening cascade further demonstrated the platform’s effectiveness in streamlining identification, referral, and notification processes (3).

While the apparent drop in diagnostic testing during the MATS period may raise concerns, it should be interpreted in light of differences in how presumptive TB cases and diagnostic evaluation were captured across periods, as well as persistent access barriers. In the pre-MATS period, presumptive cases were derived from routine programme registers and NTBLCP surveillance systems, where “presumptive TB” was often documented only when testing was initiated; consequently, most recorded presumptive cases were already in the diagnostic pathway, yielding very high testing coverage. In contrast, during the MATS period, the application captured a larger pool earlier in the care pathway at the screening stage, including symptomatic presumptive clients who did not complete referral for testing and those recorded as presumptive without TB-compatible symptoms who were intentionally excluded from testing. This widened the denominator and made downstream attrition more visible. As such, the sharp contrast between periods is unlikely to be explained solely by access barriers (which existed in both periods), but also by improved capture and classification of presumptive clients under MATS and clearer documentation of where attrition occurs along the cascade.

Beyond measurement and definitional differences, testing uptake during the MATS period was plausibly constrained by longstanding structural barriers to accessing diagnostics in Nigeria. Although TB treatment is provided free of charge after diagnosis, patients commonly bear the costs of diagnostic testing and related indirect costs such as transport and time, which likely discouraged follow-through. Health workers interviewed noted that structural and service barriers influenced linkage of presumptive clients to TB diagnosis, including low awareness of TB protocols, broader health system challenges, health workers’ attitudes, and financial constraints; stigma and distance were also frequently highlighted. Similarly, presumptive clients cited accessibility-related reasons for declining testing, including long distances to facilities, inability to afford transport fares, healthcare workers’ attitude, stigma, fear of the unknown, and cultural or religious beliefs. Taken together, the observed gap between presumptive identification and testing is therefore better interpreted as a reflection of health-system and financial access constraints, combined with earlier and more granular capture of presumptive cases under MATS, rather than a limitation of the application itself. Accordingly, when restricted to symptomatic presumptive cases eligible for testing (excluding presumptive clients with no TB-compatible symptoms), 17.9% (27,901/155,869) completed diagnostic testing during the MATS period.

Regionally, the North-West and North-Central zones demonstrated the most substantial improvements in diagnostic positivity, reflecting the tool’s adaptability to varied operational contexts (3). These regional variations underscore the importance of tailoring digital health interventions to local conditions and health system capacities. MATS’ integration across these settings proved not only feasible but also impactful, reinforcing global calls for digital innovation to close TB detection gaps (5,19).

The role of the private sector was also reinforced, with private-for-profit facilities and patent medicine vendors (PMVs) together accounting for over 62% of all presumptive TB identifications. This finding emphasises the critical importance of engaging informal and non-state actors in TB surveillance, particularly in a health system where a significant portion of care-seeking occurs outside the public sector. MATS effectively expanded the reach of TB services into these fragmented spaces, enhancing equity in case-finding coverage (6,22,26).

Beyond improving detection, MATS strengthened the architecture of TB surveillance. Health workers cited improvements in record-keeping, timeliness, and patient tracking, all of which are essential for effective public health response (3,4,11). While operational issues such as unstable internet connectivity, limited device capacity, and data upload challenges were consistently reported, these barriers reflect infrastructural gaps in the broader health system rather than deficiencies inherent to the digital tool itself (3,4,7,24).

The potential for MATS integration with broader health interventions, particularly HIV service delivery, was also noted. Although awareness of such integration was limited among some participants, others highlighted existing co-management pathways. This points to a promising opportunity for MATS to evolve into a cross-programmatic digital platform, consistent with WHO guidance on integrated care and service delivery (19,24,28).

User experiences further validated MATS’ effectiveness. Clients appreciated the efficiency, structured interactions, and reduced waiting times associated with digital screening. Health workers valued the platform’s ease of use and impact on workflow. However, persistent barriers to diagnostic follow-through—transport costs, facility wait times, stigma, and limited awareness—remain significant. These constraints are emblematic of wider health system inequities and reinforce the need for complementary investments in patient support, community sensitisation, and health system strengthening (8,18,21).

Participants across stakeholder groups provided practical recommendations for improving MATS: enhancing internet connectivity, expanding offline functionality, providing smartphones for health workers, simplifying data entry, and launching public awareness campaigns. These suggestions align with best practices for user-centred digital health design and WHO’s implementation guidelines for digital TB interventions (19). Importantly, they signal the value of continuous frontline feedback in refining health innovations.

Despite the observed decrease in diagnostic evaluation rates, the study demonstrated that MATS improved the efficiency and effectiveness of TB case detection. It also enhanced surveillance integration and offered actionable pathways for scale-up. These results affirm that a well-targeted, technology-enabled intervention can yield meaningful health system improvements when adapted to real-world settings and accompanied by broader enabling conditions (1,6,18,21).

Ultimately, MATS advances the evidence base for digital innovation in TB control, particularly within pluralistic health systems like Nigeria’s. It contributes to understanding how mobile applications can be leveraged to enhance case detection, improve surveillance, and deliver more responsive, equitable care. These findings provide a strong platform for developing policy recommendations and future research priorities that can sustain and extend the gains observed in this evaluation (3,4,23).

### Policy and Programmatic Recommendations

i. Invest in digital infrastructure: Expand reliable internet access and supply dedicated smartphones to frontline health workers to support uninterrupted MATS implementation across all facility types.
ii. Strengthen workforce capacity: Institutionalise routine training and refresher programs focused on MATS use, digital literacy, data entry, and troubleshooting to sustain implementation quality.
iii. Improve patient linkage: Introduce patient-centred incentives such as transport support and community awareness campaigns to address barriers to diagnostic follow-through and reduce TB-related stigma.
iv. Enhance application functionality: Upgrade MATS with greater offline capability, simplified user interfaces, and broader device compatibility to reduce entry delays and minimize data loss.
v. Promote integration with other health services: Align MATS with broader disease platforms, especially TB/HIV co-management, to increase efficiency and expand its utility across health system priorities.

### Limitations of the Study

Several limitations should be considered when interpreting the study’s findings. The reduction in the proportion of presumptive cases tested during MATS implementation likely reflects broader operational and health system disruptions such as logistical barriers and patient-level constraints rather than shortcomings of the application itself. Additionally, the use of facility-based data may underestimate true service uptake by excluding individuals lost to follow-up or those who sought care outside project-supported facilities. Lastly, the qualitative insights, while valuable, are subject to recall and social desirability bias, as they rely on participants’ retrospective accounts and perceptions, which may not fully capture the complexities of implementation in real time.

### Future Research Directions

Future research should focus on longitudinal evaluations to determine the sustained impact and cost-effectiveness of MATS and similar digital health tools across diverse health system settings (12,14,18). Further investigation into interventions that address the persistent barriers to patient linkages, such as stigma, transportation costs, and service delays, will be crucial for improving diagnostic follow-through. Comparative studies assessing integration with other digital platforms or health services, including TB-HIV co-management, would also provide valuable insights into scalable models for comprehensive disease control (16,17,20). These avenues of inquiry will help inform more effective and inclusive digital health strategies for TB care in Nigeria and other high-burden countries.

## Data Availability

All data produced in the present study are available upon reasonable request to the authors

## END NOTE

### Authors’ contributions

GPGP and TRdW, conceived the study and review and approved the final version of the manuscript; AS, in collaboration with tuberculosis experts, developed MATS and participated in the implementation of the tool; NN, supported the development and implementation of the tool and reviewed and approved the final version of the manuscript. BO, CO, MS, and IG supervised study implementation, oversaw data collection and analysis, and critically reviewed the manuscript. EA conducted the data analysis, while SW drafted the initial version of the manuscript. All authors reviewed and approved the final manuscript.

### Ethical considerations

(NHREC/01/01/2007-15/09/2021)

## Acknowledgments

We would like to thank the financial support of the Global Fund for the MATS intervention and from the Ministry of Foreign Affairs of the Netherlands for its evaluation.

## Notes

### Competing Interest Statement

The authors have declared no competing interest.

### Author Declarations

Approval Body: National Health Research Ethics Committee (NHREC), Nigeria Approval Number: NHREC/01/01/2007-15/09/2021

